# Internet Use impact on Physical Health during COVID-19 pandemic in Bangladesh: A Web-based Cross-sectional study

**DOI:** 10.1101/2021.08.06.21261689

**Authors:** Tanvir Abir, Uchechukwu Levi Osuagwu, Dewan Muhammad Nur–A Yazdani, Abdullah Al Mamun, Kaniz Kakon, Anas A. Salamah, Noor Raihani Zainol, Mansura Khanam, Kingsley Emwinyore Agho

**Affiliations:** International University of Business, Agriculture and Technology—IUBAT University, Dhaka 1230, Bangladesh; Diabetes, Obesity and Metabolism Translational Research Unit, Western Sydney University, Campbelltown, NSW 2560, Australia; African Vision Research Institute (AVRI), University of KwaZulu-Natal, Westville Campus, Durban, 3629, South Africa; UCSI University, Kuala Lumpur 56000, Malaysia; Department of Philosophy, College of Arts and Sciences—CAAS, International University of Business, Agriculture and Technology—IUBAT University, Dhaka 1230, Bangladesh; Department of Management Information Systems, College of Business Administration, Prince Sattam Bin Aziz University, 165 Al-Kharj 11942, Saudi Arabia; Faculty of Entrepreneurship and Business, Universiti Malaysia Kelantan, 16100 Kota Bharu, Malaysia; Nutrition and Clinical Services Division, icddr,b Bangladesh; School of Health Science, Western Sydney University, Campbelltown, NSW 2560, Australia; African Vision Research Institute (AVRI), University of KwaZulu-Natal, Westville Campus, Durban, 3629, South Africa

**Keywords:** Internet use, Coronavirus, Survey, Headache, Back pain, Neck pain, Physical health

## Abstract

**Background:** Bangladesh ranked fifth largest internet user in Asia. Past studies on internet use have focused on its impact on mental health, with little known about its impact on the physical health of individuals during COVID-19 pandemic. Hence, this study examines the impact of Internet use frequency on physical health during the Covid-19 lockdown in Bangladesh.

**Methods:** A web-based cross-sectional study on 3242 individuals aged 18 and above was conducted from 2^nd^ August – 1^st^ October 1, 2020, during the lockdown in Bangladesh. The survey covers demographics, Internet use frequency and physical health questions. Multiple linear regression analyses were used to examine the impact of internet use frequency on physical health.

**Results:** The result indicated that 72.5%, 69.9%, 65.1% and 55.3% reported headache, back pain, numbness of the fingers and neck pain, respectively. The multivariable analyses showed increased physical health impact among regular (coefficient β =0.52, 95% confidence interval [CI]: 0.18-0.85, *P*=0.003), frequent (β = 1.21, 95%CI: 0.88–1.54, *P* < .001) and intense (β = 2.24, 95%CI: 1.91–2.57, *P* < .001) internet users. Other factors associated with physical health scores were gender, income (in Taka), occupation, regions, and working status.

**Conclusion:** Frequent, intensive, and extensive use of the internet were strong predictors of increased physical health problems, and the study suggests the need for raising awareness of physical health problems triggered by high internet users among the high socioeconomic group in Bangladesh.

## INTRODUCTION

The coronavirus disease 2019 (COVID-19) pandemic and the associated public health interventions, including social distancing, home quarantine measures, and ‘stay-at-home’ orders, put in place by the various governments, led to a surge in internet usage during the pandemic ^**[1]**^. The lockdown measures resulted in widespread and unprecedented social disruption ^**[2]**^ as non-essential businesses were closed or some employees worked from home and unemployment increased ^**[3]**^. There were massive changes in internet usage patterns and user behaviour with employees having to adjust to the new “normals” - with meetings going completely online, office work shifting to the home, leading to new emerging patterns of work. These changes have come across most organizations, businesses, societies, or governments and suddenly, leaving barely any time for organizations and people to plan for, prepare and implement new setups and arrangements. Society has had to adjust, try, experiment, and find ways that did not exist before the pandemic ^**[1]**^, and these may have implications on the physical health of individuals.

People are now spending even more time with technology while consuming news media, watching television, using social media to connect with others, utilizing lifestyle apps to shop for groceries and other consumer goods, and engaging in home workouts ^**[2]**^. In Bangladesh, this kept parts of the economy going ^**[4]**^ as the country witnessed a boom in internet usage due to the fast-growing mobile internet and the government’s push for digitalization. Engaging in social media platforms is now a routine activity for children and adults, enhancing communication, social connection, and even technical skills ^**[5]**^. Platforms such as Facebook and Twitter offer multiple daily opportunities for connecting with friends, classmates, and people with shared interests. The number of people using such sites has also increased dramatically over the last five years, as shown in a recent poll ^**[6]**^. A past study reported that about 22% of teenagers log on to their favorite social media site more than ten times a day, and more than half of adolescents log on to a social media site more than once a day ^**[6**,**7]**^. Seventy-five percent of teenagers now own cell phones, 25% use them for social media, 54% use them for texting, and 24% use them for instant messaging ^**[7]**^.

According to the Bangladesh Telecommunication Regulatory Commission, there were 99,428 million (approx. 60.9% of the population) internet users in February 2020 ^**[8]**^. Though there is anecdotal early evidence that this may lead to increased productivity, it has also led to increased stress ^**[9**,**10]**^ where employees must learn new technologies, be available for work at almost all times, stay with digital devices all the time, and cope with multi-tasking ^**[1]**^. The use of video-conferencing and content delivery services such as Zoom and Akamai also increased ^**[1]**^. Those working from home using this technology have found themselves under intense scrutiny and tension ^**[11]**^.

That a large part of the peoples’ social and emotional development is occurring while on the Internet and cell phones may affect the physical health of individuals considering the long hours spent on the internet during the mandatory stay at home orders which expose the population to lots of misinformation ^**[12]**^ known to affect physical and mental health ^**[13]**^. Since this high use of information systems may become the new normal, this study examined the impact of internet usage frequency during the COVID-19 pandemic on Bangladeshi residents’ physical and mental health and determined the level of addiction to social media platforms used by the people.

### Ethical consideration

Permission for this study (IUBAT/AR/2021/001) was obtained from the Institutional Review Board of International University of Business, Agriculture and Technology (IUBAT), Dhaka, Bangladesh. The study adhered to the tenets of the Declaration of Helsinki as revised in Fortaleza. All the participants were informed about the specific objective of this study before and consent was obtained from all participants before completing the questionnaire through an online preamble. To avoid repeated responses and in order to ensure the validity of data to some extent, participants were able to complete the survey only once as data was restricted to their IP addresses and device. Participants could terminate the survey at any time they desired. Anonymity and confidentiality of the data were ensured.

## METHODS

### Study population and sampling

By convenience sampling, 3242 questionnaires were collected from adults aged 18 and over living in eight divisions across Bangladesh during the pandemic. All questionnaires were web-based, participation was voluntary, and each participant was informed about the study’s background through an online preamble. No reward or incentive was offered for participation. There may be more than one questionnaire filled by the same person that affects the statistical analysis results of questionnaire data. In order to ensure the validity of data to some extent, restricting the same IP address and device are applied to the system, which means the participant of the same IP address or device only can fill once.

### Data Collection

Data were collected using self-administered web-based questionnaires distributed to various groups of people via the E-link using numerous mailing lists and social network sites including Facebook, WhatsApp and Twitter. Due to the mandatory lockdown, no paper questionnaires were distributed, making it difficult to reach the very remote regions.

### Dependent variable

Ten items of physical complaints reported in ***Table 2*** below were used to determine the impact of individual physical health on Internet use. The respondents used a close-end response [‘Yes’ or ‘No’] to indicate whether they suffered from the following ten problems (Back and neck pain, numbness in fingers, headaches, inability to sleep, dry eyes or other vision problems, poor nutrition, poor personal hygiene, weight gain/loss and loss of appetite] during or after pro-longed Internet use. Binary scores for individual questions were summed to give a physical health score which ranged from 0-10, and the Kuder Richardson coefficient measuring internal consistency among the physical health scores ranged from 0.65 to 0.78, indicating a satisfactory level of reliability.

### Main study factor

The main study variable was the respondent’s Internet usage, including the average daily amount of time spent on the internet and dependence on networks. The frequency of internet use was categorized as ‘seldom’ (<1 hour per day), ‘casual’ (≥1 and <3 hours per day), ‘regular’ (≥3 and <5 hours per day), ‘frequent’ (≥5 and <7 hours per day), ‘intense’ (≥7 hours per day) ^**[14]**^.

### Independent variables

Independent variables used for the study analyses were based on previous studies ^**[14-16]**^. Independent variables included demographic characteristics (i.e., gender, age [categorized as 18-27, 28-37, 38 years and over], marital status [single, married, divorced/widowed], mothers level of education [Higher education, Bachelor, Intermediate], place of residence, employment status [Employed, unemployed/student], occupation [healthcare and non-healthcare worker], and income in Taka [lower, middle and high-income earners].

### Statistical Analysis

All statistical analyses were performed in Stata version 14.1 (Stata Corp. 2015, College Station USA) mainly using descriptive statistics; t-test used compares the means of two independent groups, One-way analysis of variance (one-way ANOVA) used compares the means of more than two independent groups, correlation analyses, and regression analysis. All confounding variables with a *P* < .20 were retained in the univariate linear regression analysis and were used to build a multivariable model ^**[17]**^. A manual elimination procedure was applied for multivariate linear regression analyses to remove non-significant variables (*P* > .05). The main study factor variable (Internet usage frequency) was added to all significant confounding variables after elimination processes. The main study factor and independent variables associated with physical health scores (*P* < .05) were reported. In the regression analysis, we checked for homogeneity of variance and multicollinearity, including Variance Inflation Factors (VIF) and the VIF < 4 was considered suitable ^**[18]**^:

## RESULTS

### Demographic Characteristics According to the Frequency of Internet Use among adults in Bangladesh

***Table 1*** depicts the main demographic characteristics, including gender, education and age group on the five groups extracted. It is worth noting that the education in this paper refers to that of the mothers’ maximum educational level. The majority of participants were males (1260, 61.1%), married (2632, 81.2%) and employed (2864, 88.3%) in non-healthcare sectors (2567, 79.2%), and for many, their mothers had completed a bachelor degree or higher. About half of the respondents earned more than 70,000 Taka at the time of data collection (***Table 1***).

**Table 1.**
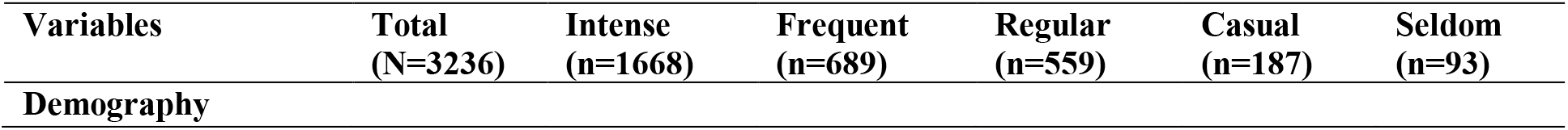
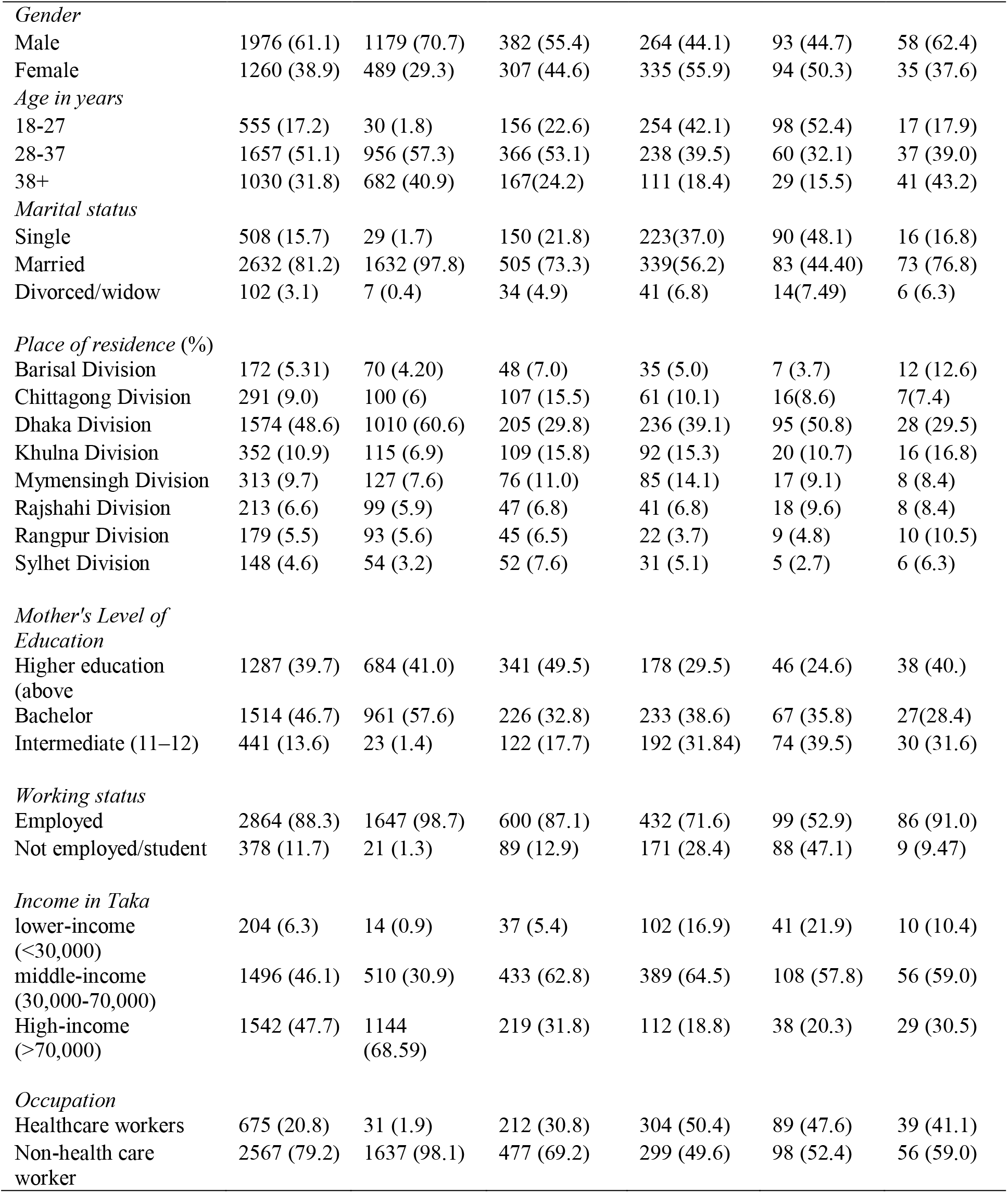
Demographic Characteristics According to the Frequency of Internet Use among adults in Bangladesh. Values are expressed as numbers and percentages (n, %).

About two-thirds of the respondents that reported intense use of the internet were males, while females were more likely to report regular and casual use of the internet. More than half of intense and frequent internet users were aged 28-37years, and a similar proportion of casual internet users were younger (18-27years). Nearly all intense internet users were employed and working in the non-Health care sector during the study.

### Prevalence and 95% confidence intervals of physical complaints by gender among internet users

***Table 2*** depicts the prevalence and 95%CIs of the adverse effects on individual physical health complaints examined in this study in males and females. Headache 72.50% [95%CI 70.93%, 74.01%] was the prevalent complaint of the participants, while 69.87% [68.27%, 71.43%] and 65.10% [63.42%, 66.71%] complained of Back pain and numbness of the fingers, respectively. Males reported a higher prevalence of back pain, numbness of the finger, headaches, and neck pains than females as the confidence intervals do not overlap (see ***Table 2***). By contrast, females reported a higher prevalence of poor nutrition, poor personal hygiene, and loss of appetite compared with men (***Table 2***).

**Table 2.**
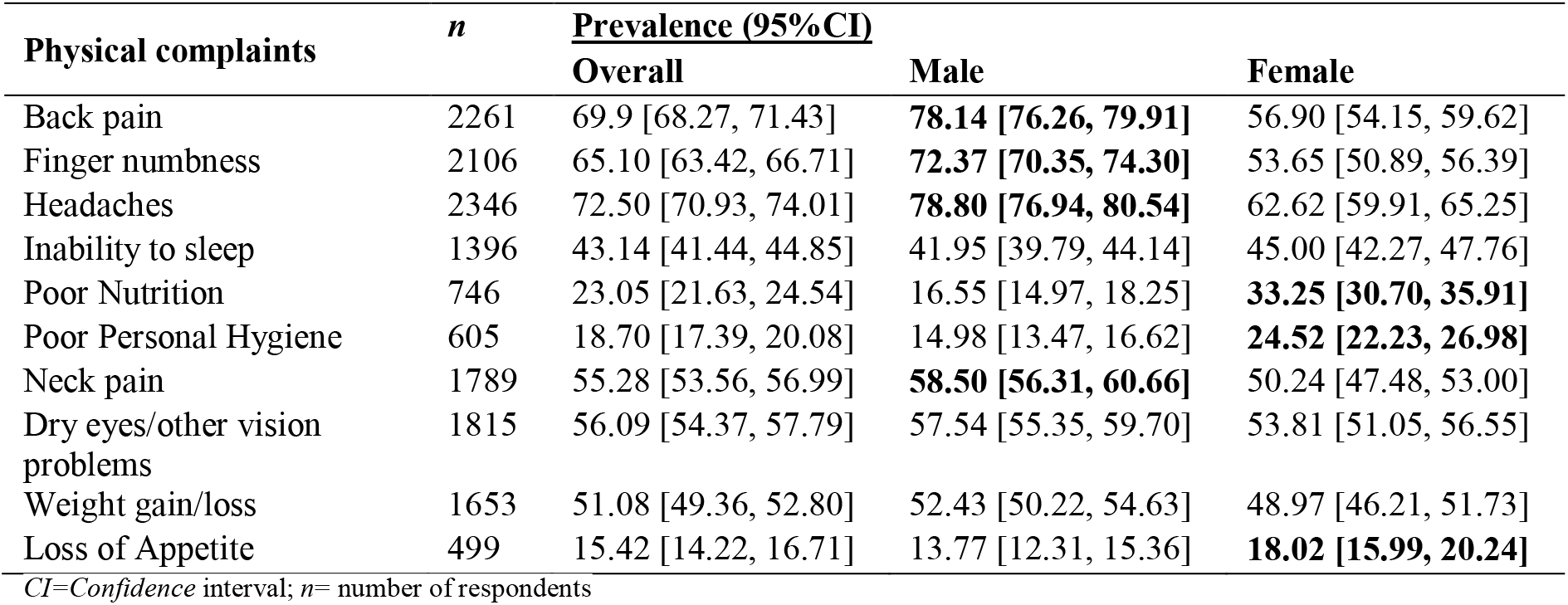
Gender differences in the prevalence of physical health scores among internet users in Bangladesh. *Significant variables between males and females are bolded because the confidence intervals did not overlap*.

| Physical complaints | n | Prevalence (95% CI) |  |  |
| --- | --- | --- | --- | --- |
|  |  | Overall | Male | Female |
| Back pain | 2261 | 69.9 [68.27, 71.43] | **78.14 [76.26, 79.91]** | 56.90 [54.15, 59.62] |
| Finger numbness | 2106 | 65.10 [63.42, 66.71] | **72.37 [70.35, 74.30]** | 53.65 [50.89, 56.39] |
| Headaches | 2346 | 72.50 [70.93, 74.01] | **78.80 [76.94, 80.54]** | 62.62 [59.91, 65.25] |
| Inability to sleep | 1396 | 43.14 [41.44, 44.85] | 41.95 [39.79, 44.14] | 45.00 [42.27, 47.76] |
| Poor Nutrition | 746 | 23.05 [21.63, 24.54] | 16.55 [14.97, 18.25] | **33.25 [30.70, 35.91]** |
| Poor Personal Hygiene | 605 | 18.70 [17.39, 20.08] | 14.98 [13.47, 16.62] | **24.52 [22.23, 26.98]** |
| Neck pain | 1789 | 55.28 [53.56, 56.99] | **58.50 [56.31, 60.66]** | 50.24 [47.48, 53.00] |
| Dry eyes/other vision problems | 1815 | 56.09 [54.37, 57.79] | 57.54 [55.35, 59.70] | 53.81 [51.05, 56.55] |
| Weight gain/loss | 1653 | 51.08 [49.36, 52.80] | 52.43 [50.22, 54.63] | 48.97 [46.21, 51.73] |
| Loss of Appetite | 499 | 15.42 [14.22, 16.71] | 13.77 [12.31, 15.36] | **18.02 [15.99, 20.24]** |

CI=Confidence interval; n= number of respondents

***Figure 1*** shows the mean scores of physical health by internet use frequency in males and females. From the figure, the mean scores of physical health by intensive internet use was 5.6 for males and 5.4 for females. This physical health mean scores could be translated as percentage mean scores of males (56%) and females (54%), respectively. Physical health mean scores were lower among the casual user of internet (3.2 for females and 3.0 for males) than seldom users of internet (3.1 for females and 3.4 for males)

**Figure 1.**
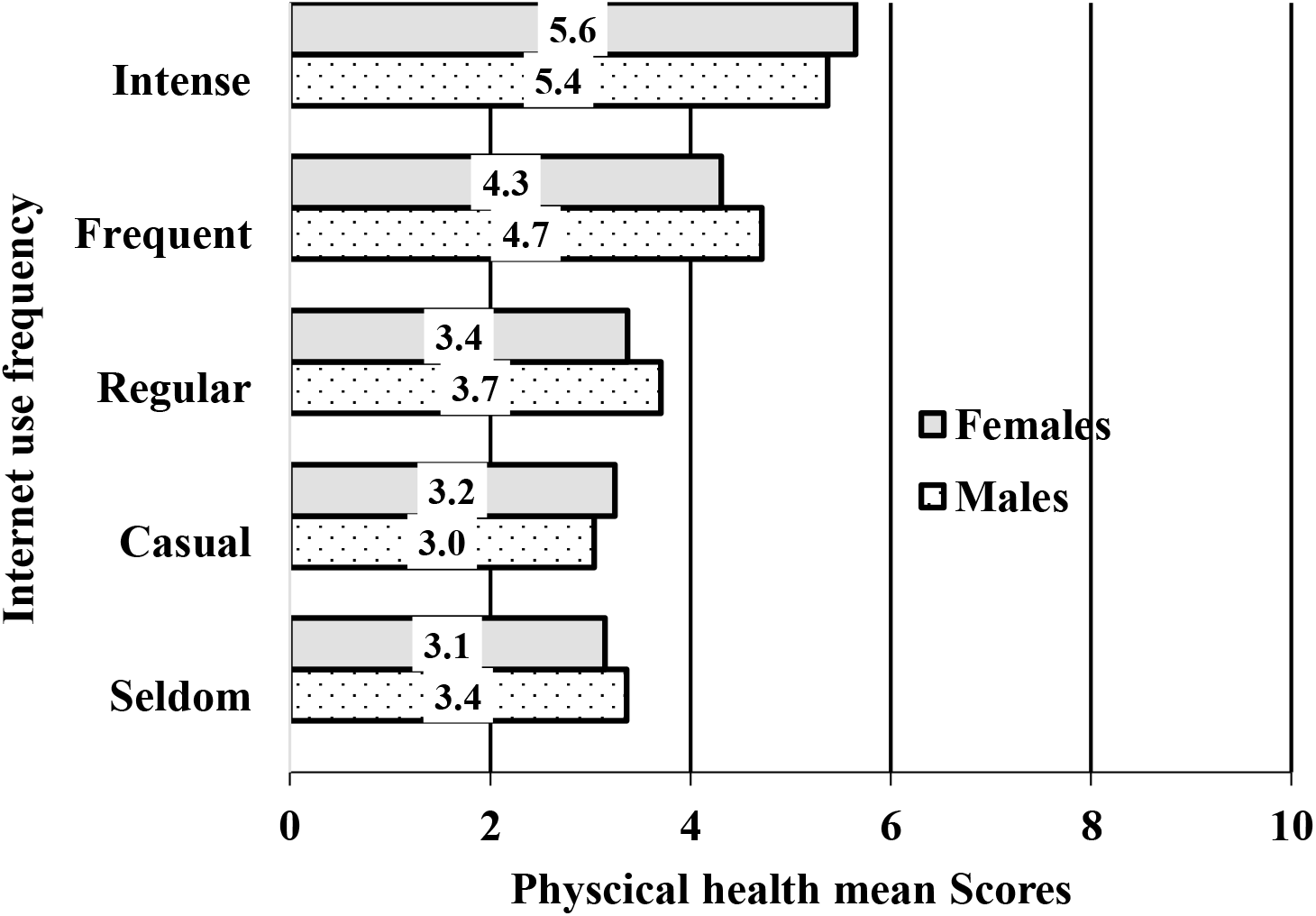
Mean scores by gender of the physical health by internet use frequency in Bangladesh

### Unadjusted Analysis for the association between prolonged internet use and physical health complaints

Table 3 shows the mean scores of physical health complaints and the unadjusted coefficients for the associations with the sociodemographic variables. From the table, the mean scores for physical health complaints varied across variables. The unadjusted coefficients revealed significantly higher physical health problems due to prolonged internet use in those aged

**Table 3:**
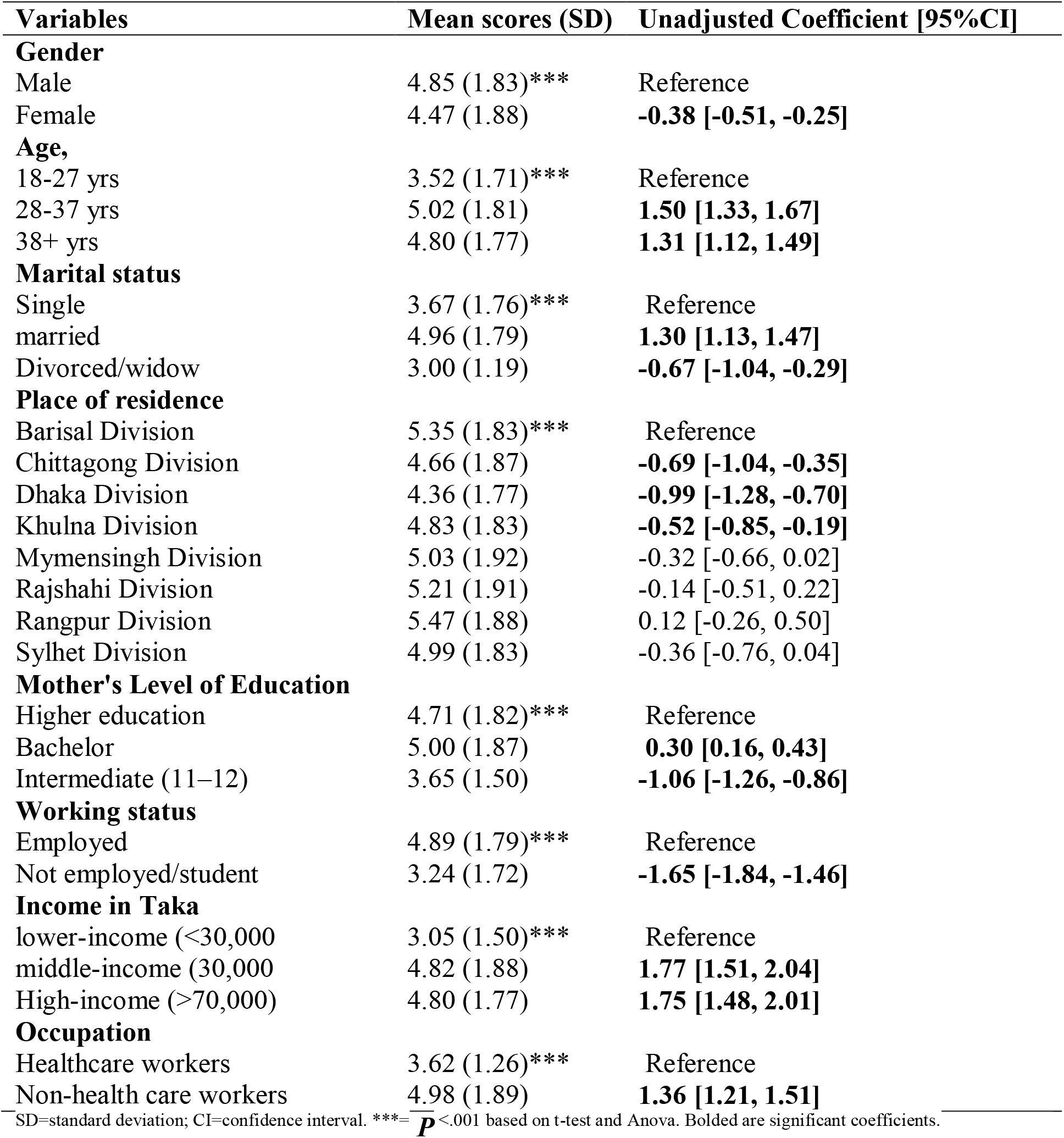
**Mean scores and** Univariate linear regression of Physical Complaints among Internet Users.

| Variables | Mean scores (SD) | Unadjusted Coefficient [95%CI] |
| --- | --- | --- |
| **Gender** |  |  |
| Male | 4.85 (1.83)*** | Reference |
| Female | 4.47 (1.88) | **-0.38 [-0.51, -0.25]** |
| **Age,** |  |  |
| 18-27 yrs | 3.52 (1.71)*** | Reference |
| 28-37 yrs | 5.02 (1.81) | **1.50 [1.33, 1.67]** |
| 38+ yrs | 4.80 (1.77) | **1.31 [1.12, 1.49]** |
| **Marital status** |  |  |
| Single | 3.67 (1.76)*** | Reference |
| married | 4.96 (1.79) | **1.30 [1.13, 1.47]** |
| Divorced/widow | 3.00 (1.19) | **-0.67 [-1.04, -0.29]** |
| **Place of residence** |  |  |
| Barisal Division | 5.35 (1.83)*** | Reference |
| Chittagong Division | 4.66 (1.87) | **-0.69 [-1.04, -0.35]** |
| Dhaka Division | 4.36 (1.77) | **-0.99 [-1.28, -0.70]** |
| Khulna Division | 4.83 (1.83) | **-0.52 [-0.85, -0.19]** |
| Mymensingh Division | 5.03 (1.92) | -0.32 [-0.66, 0.02] |
| Rajshahi Division | 5.21 (1.91) | -0.14 [-0.51, 0.22] |
| Rangpur Division | 5.47 (1.88) | 0.12 [-0.26, 0.50] |
| Sylhet Division | 4.99 (1.83) | -0.36 [-0.76, 0.04] |
| **Mother's Level of Education** |  |  |
| Higher education | 4.71 (1.82)*** | Reference |
| Bachelor | 5.00 (1.87) | **0.30 [0.16, 0.43]** |
| Intermediate (11–12) | 3.65 (1.50) | **-1.06 [-1.26, -0.86]** |
| **Working status** |  |  |
| Employed | 4.89 (1.79)*** | Reference |
| Not employed/student | 3.24 (1.72) | **-1.65 [-1.84, -1.46]** |
| **Income in Taka** |  |  |
| lower-income (<30,000) | 3.05 (1.50)*** | Reference |
| middle-income (30,000) | 4.82 (1.88) | **1.77 [1.51, 2.04]** |
| High-income (>70,000) | 4.80 (1.77) | **1.75 [1.48, 2.01]** |
| **Occupation** |  |  |
| Healthcare workers | 3.62 (1.26)*** | Reference |
| Non-health care workers | 4.98 (1.89) | **1.36 [1.21, 1.51]** |

SD=standard deviation; CI=confidence interval. \*\*\*= $P < .001$ based on t-test and Anova. Bolded are significant coefficients.

>28years, married, completed bachelor education, middle-high income earners and non-healthcare workers compared with other groups. In contrast, females, those who were divorced or widowed, those with intermediate education, non-workers/students and respondents who resided in Chittagong, Dhaka and Khulna during the study period had significantly lower mean scores of physical health complaints from prolonged internet usage compared with the other groups.

### The impact of the frequency of internet use on physical health scores

***Figure 2*** presents the unadjusted and adjusted odds ratio and their confidence intervals for the association between frequency of internet use and physical health symptoms during the COVID-19. In the unadjusted analysis, frequent and intensive internet users during the COVID-19 significantly reported higher physical health problems than seldom users of the internet. After adjusting for the independent variables used in this study, regular, frequent, and intensive internet users during the COVID-19 had significantly higher physical health problems than those who seldom used the internet (see ***Figure 2***).

**Figure 2:**
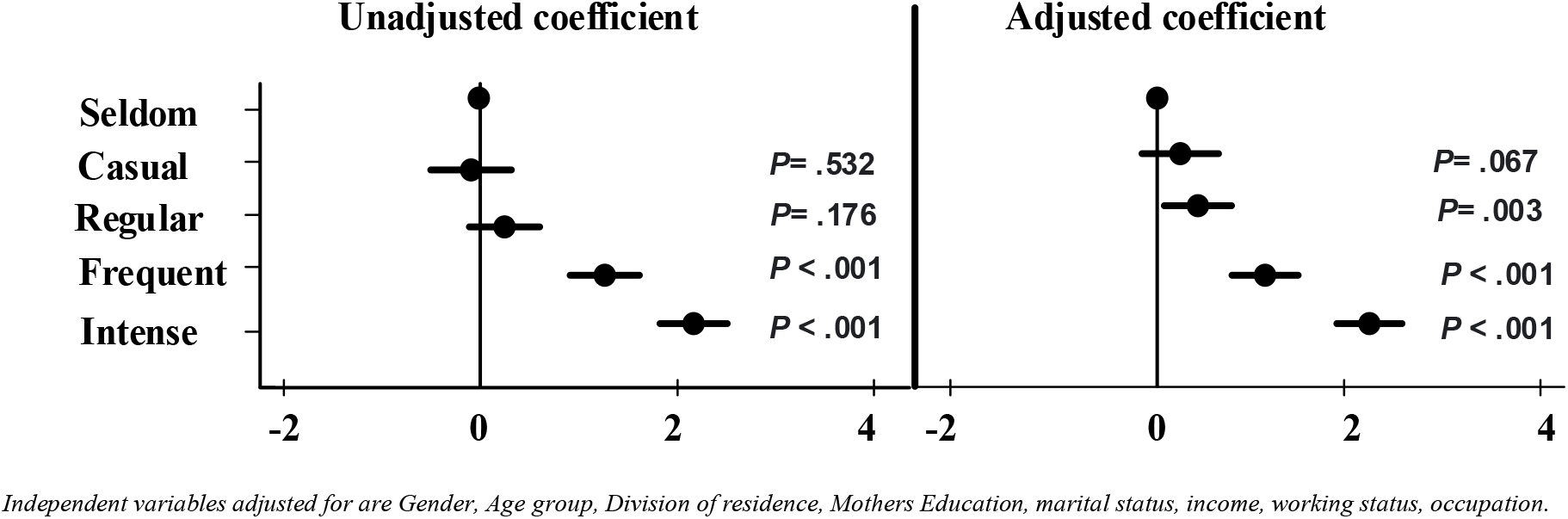
The impact of frequency of internet use on physical health scores: Unadjusted and adjusted coefficient and their 95% confidence intervals.

Marital status, age, gender, place of residence, level of education, working status, occupation, and income level were all associated with the physical health complaints among internet users in Bangladesh (***Supplementary Table S1 for details***).

## DISCUSSION

A considerable body of literature has emerged over the past two decades assessing the relationship between problematic or addictive (excessive) use of the internet and individuals’ psychological/mental well-being. Comparatively, very little research has evaluated the relationship between various internet use frequencies and the individual’s physical health. The COVID-19 pandemic caused a drastic surge in the use of the internet globally and Bangladesh ^**[1]**^, and this cross-sectional study investigated the relationship between the frequency of internet use and the physical health of Bangladeshi respondents. The findings demonstrated that more than one in every two persons in this study used the internet for seven or more hours each day. This was associated with 2.2 folds increase in physical health problems compared with seldom users (<1 hour per day). Headache, back pain and numbness of the fingers (reported by over seventy percent of internet users) were the predominant complaints. There was a positive association between physical health problems and the number of daily hours spent on the internet. After adjusting for the frequency of internet use, we found that middle to high-income earners and those working in non-healthcare sectors experienced more significant physical health problems due to internet use. In comparison, students and widowed or divorced persons reported fewer physical health problems compared with other respondents.

This study found that headache was the most prevalent symptom reported by internet users in Bangladesh, followed by back pain and numbness of the fingers. These complaints were significantly increased among frequent internet users and intense users, which agrees with the findings of significant association between extensive use of the internet and physical health problems in previous studies ^**[19-22]**^. A study in Spain reported that headache was the most common complaint among internet users during the COVID-19 pandemic ^**[22]**^. Similarly, Cao et al.s study (2009), after examining adolescents, found that those who used the internet excessively were more likely to report psychosomatic symptoms including lack of physical energy, physiological dysfunction, weakened immunity, emotional symptoms, behavioural symptoms and social adaptation problems and had lower life satisfaction scores compared with normal internet users ^**[20]**^.

There are also reports that school children and other graduate students in China ^**[23]**^ and Italy ^**[24]**^ experienced increased psychological distress following excessive internet use during the COVID-19 pandemic. Such addiction to the internet can result in the problem of self-care, difficulty in performing daily routine, and mental health effects of anxiety and depression ^**[25]**^. The negative effects of internet addiction on health may be related to the alterations in the individuals neuroanatomical and neurochemical mechanisms, including cortical thinning of various components of the brain and dopaminergic reward circuitry, during excessive use of the internet ^**[25]**^. While it is imperative that such behaviours, especially during a pandemic, remain at a moderate and regulated level, identifying the population at risk of the problematic use of the internet is necessary in order to prevent the associated mental health problems ^**[26]**^ and improve work or school functions ^**[27]**^.

The present study further revealed that, compared with women, a higher proportion of men used the internet excessively during the pandemic and thus experienced more significant physical health problems. However, this association was significant only when it interacted with the frequency of internet use. Previous studies among Taiwanese residents ^**[28]**^ and those in mainland China ^**[29]**^ found that male gender was a potential risk factor for internet addiction among adolescents. This association of gender and physical health complaints found in the present study could be explained by the patriarchal nature of the Bangladesh society, where women and girls rarely go outside of their homes even before the pandemic ^**[30]**^ but preferred to stay home and spend more time on the Internet. Whereas these women may have developed adaptive strategies, it was a different experience for the male counterpart, which were used to staying outside/working prior to the lockdown. For these men, the lockdown measures and transition to work online and/or staying at home required them to adjust quickly, try, experiment, and find ways that did not exist before the pandemic ^**[1]**^ as educational institutions, businesses and workplaces were shut down for such a long duration ^**[31]**^. These adjustments may have affected their physical health.

Another finding of this study was the significant relationship between income class and occupational status (non-healthcare worker) with the physical health problems experienced by internet users. Earning above 30,000 Taka was associated with higher physical health problems due to internet usage after adjusting for the frequency of internet use. In line with this finding, Islam et al ^**[32]**^ on the correlation between problematic (excessive) internet use (PIU) and lifestyle during the COVID-19 pandemic using data from a large sample of Bangladeshi youth and adults (n=13,525) found that younger age, higher education, cigarette smoking, more sleep, less physical activity, gaming and social media use were associated with PIU ^**[32]**^. This research has depicted the physical health concerns related to prolonged internet usage during the COVID-19 pandemic. A similar association was found in this research between educational status, age and physical health complaints among internet users in Bangladesh; however, these associations were dependent on the frequency of internet use, which was not assessed in the previous study ^**[32]**^.

The finding that non-healthcare workers had higher physical health problems compared with health care workers in this study is consistent with the finding of poorer psychological well-being among non-health-care workers who used the Internet to obtain information on COVID-19 compared with those who received COVID-19 information from medical staff in health care settings ^**[33]**^. Although this research were unable to identify whether the non-healthcare workers in our study used the internet for COVID-19 information retrieval, it has been shown that the influx of misinformation around COVID-19 via online platforms had adverse effects on people’s health ^**[33]**^. Such problematic internet use (PIU) can lead to clinical impairment, distress ^**[34]**^ and/or deterioration in financial, familial, social, educational and/or occupational domains ^**[35]**^.

This study has some limitations. First, the cross-sectional design of this study does not allow for causality or the direction of relationships to be determined. However, appropriate analysis of cross-sectional data represents a useful initial step in identifying associations between extensive internet usage and the physical health condition of the participants. Second, as the responses were self-reported, it was not possible to verify the participants’ physical health complaints, which may lead to a response bias. Multiple assessments, interviews, and informants may have provided a richer and more thorough understanding of this topic. Third, since the survey was administered during COVID-19 lockdown using an online platform, it is possible that some respondents, especially those living in rural areas with limited internet access or people addicted to other online platforms such as gaming or social media, may not have participated in this survey. Although this findings may not represent the opinion of all internet users, a face-to-face survey was not possible at the time due to the lockdown measures. Despite these limitations, this was the first study to investigate the associations between extensive internet use and physical health symptoms in a sample of adolescents in mainland Bangladesh. Future studies could address nesting follow-ups in existing random samples that could provide timely access to longitudinal population-representative data.

## Conclusions

In summary, the study demonstrated that uncontrolled use of the internet is prevalent among the respondents in this study as more than two-third reported excessive use of the internet during the lockdown, and this doubled the adverse effects on their physical health compared with seldom users. Headaches, back pain, numbness of the fingers and neck pain were predominant among internet users as spending 3 hours or more each day on the internet led to a significant increase in the level of adverse physical health problems. The positive associations between physical complaints and non-healthcare workers, middle to high-income earners, indicate the target group for interventions to minimize this growing epidemic. The health department of the Bangladesh government has now recognized extensive internet usage as a serious public health problem. Arguably, it is time for the World Health Organization and health departments worldwide to develop effective health policies to increase public awareness of extensive internet use, especially during the pandemic. This has the potential to affect millions of children and adults and, eventually the societies and economies.

## Data Availability

Our data are included in the manuscript and raw data can be released at reasonable request.

---

**Supplementary Table 1.** Results of Multiple Regression Analysis in Prediction of Level of Physical Complaints Among Internet Users. *Bold are significant variables*.

| Variables | Adjusted Coefficients [95%CI] | P-value |
| --- | --- | --- |
| **Gender** |  |  |
| Male | Reference |  |
| Female | -0.08 [-0.20, 0.04] | 0.179 |
| **Age groups** |  |  |
| 18-27 yrs | Reference |  |
| 28-37 yrs | 0.09 [-0.13, 0.30] | 0.440 |
| 38+ yrs | -0.03 [-0.29, 0.23] | 0.818 |
| **Marital status** |  |  |
| Single | Reference |  |
| married | -0.08[-0.29, 0.12] | 0.431 |
| Divorced/widow | **-1.09[-1.44, -0.74]** | <0.0005 |
| **Place of residence** |  |  |
| Barisal Division | Reference |  |
| Chittagong Division | **-0.70 [-0.99, -0.41]** | <0.0005 |
| Dhaka Division | **-1.27 [-1.51, -1.02]** | <0.0005 |
| Khulna Division | **-0.42 [-0.70, -0.14]** | 0.003 |
| Mymensingh Division | **-0.31 [-0.59, -0.03]** | 0.031 |
| Rajshahi Division | -0.29 [-0.60, 0.01] | 0.060 |
| Rangpur Division | -0.11[-0.43, 0.21] | 0.487 |
| Sylhet Division | **-0.40[-0.74, -0.07]** | 0.018 |
| **Mother's Level of Education** |  |  |
| Higher education | Reference |  |
| Bachelor | -0.00 [-0.16, 0.16] | 0.976 |
| Intermediate (11–12) | 0.01 [-0.19, 0.21] | 0.924 |
| **Working status** |  |  |
| Employed | Reference |  |
| Not employed/student | **-0.49 [-0.72, -0.26]** | <0.0005 |
| **Income in Taka** |  |  |
| lower-income (<30,000) | Reference |  |
| middle-income (30,000-70,000) | **0.72 [0.48, 0.96]** | <0.0005 |
| High-income (>70,000) | 0.26 [-0.00, 0.53] | 0.052 |
| **Occupation** |  |  |
| Healthcare workers | Reference |  |
| Non-health care worker | **0.62 [0.47, 0.78]** | <0.0005 |
| **Internet use Frequency** |  |  |
| Seldom | Reference | - |
| Casual | 0.36 [-0.02, 0.74] | 0.067 |
| Regular | **0.52 [0.18, 0.85]** | 0.003 |
| Frequent | **1.21 [0.88, 1.54]** | <0.0005 |
| Intense | **2.24 [1.91, 2.57]** | <0.0005 |
| **Constant** | 3.18 [2.69, 3.67] | <0.0005 |

<sup>—</sup>The final model included Gender, Age group, Division, Mother's education, Income status, Employment status, Occupational status, Marital status and Internet use frequency.

